# Physical activity predicts fitness and walking capacity after stroke: a diagnostic accuracy study

**DOI:** 10.1101/2025.03.06.25323508

**Authors:** Kevin Moncion, Lynden Rodrigues, Bernat De Las Heras, Elise Wiley, Kenneth S Noguchi, Janice J Eng, Ada Tang, Marc Roig

**Affiliations:** School of Physical & Occupational Therapy, Faculty of Medicine, McGill University, Montreal, Canada; Memory and Motor Rehabilitation Laboratory (MEMORY-LAB), Feil and Oberfeld Research Centre, Jewish Rehabilitation Hospital, Montreal Center for Interdisciplinary Research in Rehabilitation (CRIR), Laval, Canada; Department of Occupational Sciences and Occupational Therapy, Faculty of Medicine, University of British Columbia, Kelowna, Canada; Centre for Chronic Disease Prevention and Management, Faculty of Medicine, University of British Columbia, Kelowna, Canada; School of Health and Exercise Sciences, Faculty of Health and Social Development, University of British Columbia, Kelowna, Canada; Department of Physical Therapy, University of British Columba and Centre for Aging SMART at Vancouver Coastal Health, Canada; School of Rehabilitation Sciences, Faculty of Health Sciences, McMaster University, Hamilton, Canada

**Keywords:** Cardiorespiratory fitness, physical activity, stroke, mobility

## Abstract

**Background and Purpose:** Clinicians need access to accurate self-reported tools that can assist with screening individuals who are at risk for fitness (V̇O_2_peak) and walking impairments (e.g., 6-minute walk test [6MWT] distance) post-stroke. The associations and diagnostic metrics between self-reported physical activity as measured by the Physical Activity Scale for People with Disabilities (PASIPD, MET-hours/week) and V̇O_2_peak (≥ 15 mL/kg/min) and 6MWT (≥ 350 m) among ≥ 6 months post-stroke were evaluated.

**Methods:** This is a secondary analysis from an aerobic exercise RCT. Participants’ age, sex, V̇O_2_peak, 6MWT distance and the PASIPD were collected at baseline. Multivariable logistic regression analyses evaluated the association between V̇O_2_peak (≥ 15 mL/kg/min), 6MWT (≥ 350 m) and the PASIPD (MET-hours/week). Predicted classifications and the Youden index identified cut points of the PASIPD.

**Results:** Eighty-five participants (n=53 males, aged 65.1 ± 9.5 years, 1.8 ± 1.2 years post-stroke) were included. Significant associations were found whereby one-unit increase in the PASIPD (MET-hours/week) was associated with a 21% increase in the odds of having a V̇O_2_peak ≥ 15 mL/kg/min [aOR = 1.21; 95% CI 1.07, 1.36, p=0.002] and 14% increase in the odds of having a 6MWT ≥ 350 m [aOR = 1.14; 95% CI 1.05, 1.23, p=0.001] with excellent area under the curve values (AUC: 0.82-0.93). Youden-derived PASIPD cut points of 8.9 and 10.6 MET-hours/week may identify individuals with fitness (≥ 15 mL/kg/min) and walking impairments (≥ 350 m)post-stroke (AUC: 0.69-0.71).

**Discussion and Conclusions:** Clinicians may use the self-reported PASIPD to identify individuals with fitness and walking impairments post-stroke.

## Introduction

A critical aspect of stroke secondary prevention is managing cardiovascular risk factors and promoting healthy lifestyles. Higher physical activity (PA) levels and cardiorespiratory fitness (V̇O_2_peak) are associated with lower all-cause mortality and lower incidence of stroke.^1–3^ Structured aerobic exercise (AE) and PA has the potential to simultaneously improve cardiorespiratory,^4^ metabolic, and neural recovery biomarkers,^5,6^ and as such, are recommended aspects of stroke rehabilitation.^7,8^

There is an interaction between V̇O_2_peak impairments, activity limitations (e.g., walking capacity), and physical inactivity post-stroke.^9^ These constructs can be assessed via “gold-standard” assessments (e.g., cardiopulmonary exercise testing [CPET],^8,10^ the 6-minute walk test [6MWT]), or accelerometry, respectively).^11^ However, these assessments are often costly and time-intensive to guide AE prescription in clinical practice.^11,12^ Indeed, stroke clinicians require access to accurate and cost-effective tools that can assist with screening and AE prescription post-stroke.^12,13^

The Physical Activity Scale for People with Disabilities (PASIPD)^14^ is a reliable self-reported tool that may be used to estimate the metabolic equivalents spent in PA (MET-hours/week) after stroke.^15^ There is currently limited evidence on whether this tool is associated with gold standard assessments of cardiorespiratory fitness or walking capacity post-stroke.^11,16,17^ However, the PASIPD may be a cost-effective screening tool that can be used to facilitate the early stratification of individuals who may benefit from more resource-intensive CPET or targeted interventions aimed at improving fitness and reducing subsequent stroke risk.

Therefore, the primary objective of this study was to examine whether the PASIPD (MET-hours/week)^14^, is associated with V̇O_2_peak (mL/kg/min) and walking capacity (6MWT distance, meters) among individuals at least 6 months post-stroke. It is hypothesized that the PASIPD will be positively and moderately associated with V̇O_2_peak,^16^ but not 6MWT.^17^ The secondary objective was to evaluate the diagnostic accuracy of the PASIPD in identifying V̇O_2_peak and 6MWT impairments post-stroke. A tertiary objective was to determine optimal PASIPD cut points that maximize the combined sensitivity and specificity for classifying individuals with V̇O_2_peak and 6MWT impairments. For each objective, V̇O_2_peak and 6MWT cut points were set as 15 mL/kg/min and 350 m, respectively, as these reference values are associated with cardiovascular disease, disability and loss of independence after stroke.^1–3,18,19^

## Methods

This study was a retrospective cross-sectional analysis of baseline data collected from a multi-site RCT (<u>NCT03614585</u>).^20,21^ Research ethics approval was obtained (HIREB 4713, CRIR-1310-0218) and informed written consent was obtained from all participants. Sample size estimation and methodological details for the RCT is reported elsewhere.^20–22^ The reporting of this study followed the STARD 2015 guidelines for diagnostic accuracy studies (Appendix 1).^23^

### Participants

Participants were recruited from the Jewish Rehabilitation Hospital (Laval, Quebec) and community members within the Montreal Metropolitan area and the Ontario Central South Stroke Network (Hamilton, Ontario). Enrollment occurred between January 2019 and August 2023. Participants were eligible if they were: between 40–80 years old and 6-60 months following the first-ever stroke confirmed by MRI/CT, living in the community, able to independently walk with or without a gait aid for at least 10 meters. Individuals were excluded from this study if they: had a stroke of non-cardiogenic origin or tumor, scored > 2 on the modified Rankin Scale,^24^ had contraindications to CPET or classified as class C or D American Heart Association Risk Criteria,^25^ were actively engaged in stroke rehabilitation services, had other neurological or musculoskeletal comorbidities, exhibited pain that worsened with exercise, or cognitive, communication, or behavioural issues that could limit their ability to provide consent or follow instructions.

### Assessments

Demographic information, including age, biological sex, and medical history were collected for all participants at baseline. All assessments took place within one week. Information regarding timing post-stroke, stroke type, and stroke severity as assessed by the National Institutes of Health Stroke Severity Scale^26^ were collected. Global cognitive function was assessed by the Montreal Cognitive Assessment.^27^ Gait speed was evaluated as a participant’s usual gait speed over a 10-meter distance using their usual gait aid as appropriate.

### Self-Reported Physical Activity

The PASIPD was considered the index questionnaire and primary independent variable of interest.^14^ The PASIPD is a 13-item questionnaire that captures information relevant to leisure time, household and work-related activity and exercise behaviours in the last 7 days.^14^ The PASIPD is scored as the hours of metabolic equivalents per week (MET-hours/week), where higher scores indicate more PA.^14^ The PASIPD has good test-retest reliability in chronic stroke^15^ and moderate construct validity in other neurological populations.^28,29^ The PASIPD was analyzed using validated instructions after all clinical assessments were completed.^14^ Since data was collected at baseline, clinical interventions were not delivered between the administration of the PASIPD (i.e., index test) and the reference standards (i.e. V̇O_2_peak and 6MWT).

### Cardiorespiratory fitness

The V̇O_2_peak, the gold standard physiological assessment of fitness,^30^ was the primary dependent variable and reference standard of interest. All participants underwent a symptom-limited CPET at baseline on recumbent stepper (NuStep T4r, NuStep LLC, Ann Arbor, MI, United States), following a protocol validated for individuals with stroke.^31^ V̇O_2_peak was collected continuously via a fitted facemask connected to a calibrated metabolic cart (Quark CPET, COSMED SrI, Rome, Italy or VMAX Encore Metabolic Cart, Sensormedic, San Diego, California). Additional CPET details are described elsewhere.^20–22^

### 6-Minute Walk Test (6MWT)

The 6MWT was the secondary dependent variable of interest and was defined as the secondary reference standard for walking capacity. The 6MWT was conducted in a 20-meter indoor hallway at baseline, and participants were instructed to walk as far as possible in 6 minutes. The total distance walked in meters was recorded after the test. Participants were permitted to use gait aids and to take rest breaks if needed. Standardized guidelines were followed during the 6MWT.^32^ The 6MWT has strong test-retest reliability and construct validity in individuals post-stroke.^33,34^

### Statistical Analyses

Participant demographic statistics were summarized as means ± standard deviation or medians (interquartile range [IQR]) for normally and non-normally distributed data, respectively, and frequencies (n, %) for categorical data. To promote sex-based reporting in exercise stroke research,^35^ baseline characteristics were disaggregated by sex. Differences in baseline characteristics between males and females were determined using independent t-tests, Wilcoxon rank-sum tests and chi-squared tests (χ^2^) for parametric, non-parametric and categorical data, respectively.

Separate multivariable logistic regression analyses were performed to examine the association between V̇O_2_peak (primary dichotomous dependent variable at a cut point of 15 mL/kg/min^36^), 6MWT distance (secondary dichotomous dependent variable at a cut point of 350m^18,19^) and the PASIPD (primary independent continuous variable measured in MET-hours/week). These cut points were pre-specified and selected since they are often referenced as cut points that are associated with cardiovascular disease, disability and loss of independence post-stroke.^1–3,18,19^ Multivariable models were adjusted for covariates of age (years, continuous), sex (male/female, binary), and the 10-m usual gait speed (m/s, continuous). The 6MWT test models did not include 10-m usual gait speed as a covariate due to collinearity. Covariate selection were driven by theory given their known influence on the primary and secondary dependent variables.^21,37,38^ Requisite assumptions, including normality, residual and influencing outliers (e.g., Cook’s distance > 4/n) and influential covariate patterns, goodness of fit (e.g., Hosmer Lemeshow), and collinearity (e.g., variance inflation factor), were assessed. Predicted classifications and diagnostic metrics were determined and reported using a probability cutoff of 0.5 for each V̇O_2_peak and 6MWT multivariable logistic regression models.

The Youden index was then used to determine the optimal PASIPD cut point that can be used to discriminate between V̇O_2_peak and 6MWT impairments by identifying the optimal cut point that maximizes the sum of sensitivity and specificity - 1.^39^ Consistent with the multivariable logistic regression models, V̇O_2_peak and 6MWT was dichotomized using cut points of 15 mL/kg/min and or 350 m, respectively. The Youden-derived PASIPD cut point was dichotomized to enable receiver operating characteristic (ROC) curve analyses and 2 × 2 tables. Pre-planned sensitivity analyses were conducted using V̇O_2_peak cut points at 12 mL/kg/min and 18 mL/kg/min, and 6MWT cut points at 280 meters, and 380 meters for the multivariable logistic regression models and Youden-derived PASIPD cut points given known associations with cardiovascular disease, disability and loss of independence post-stroke.^1–3,18,19^

The area under the curve (AUC) and it’s precision estimate (95% CI) were used to determine the overall diagnostic accuracy of the multivariable logistic regression models and the Youden-derived unadjusted PASIPD cut points, where AUC equals 0.5 when the ROC curve corresponds to random chance and 1.0 for perfect accuracy.^40^ AUC values are generally interpreted as no discrimination (AUC = 0.5), acceptable (AUC = 0.7 to 0.8), excellent (0.8 to 0.9), and perfect (0.9 to 1.0).^41^ The sensitivity, specificity, positive predictive values, negative predictive values, positive likelihood ratios, and negative likelihood ratios along with their precision estimates (95% CI) were also reported. The level of significance was set to *p* = 0.05.

All statistical analyses were conducted with STATA 16 and with the epiR package^42^ via R version 4.4.2.

## Results

In total, 85 participants were deemed eligible had complete V̇O_2_peak, 6MWT and PASIPD data at baseline (Figure 1). Participant characteristics for the entire sample and disaggregated by sex are described in Table 1.

**Figure 1.**
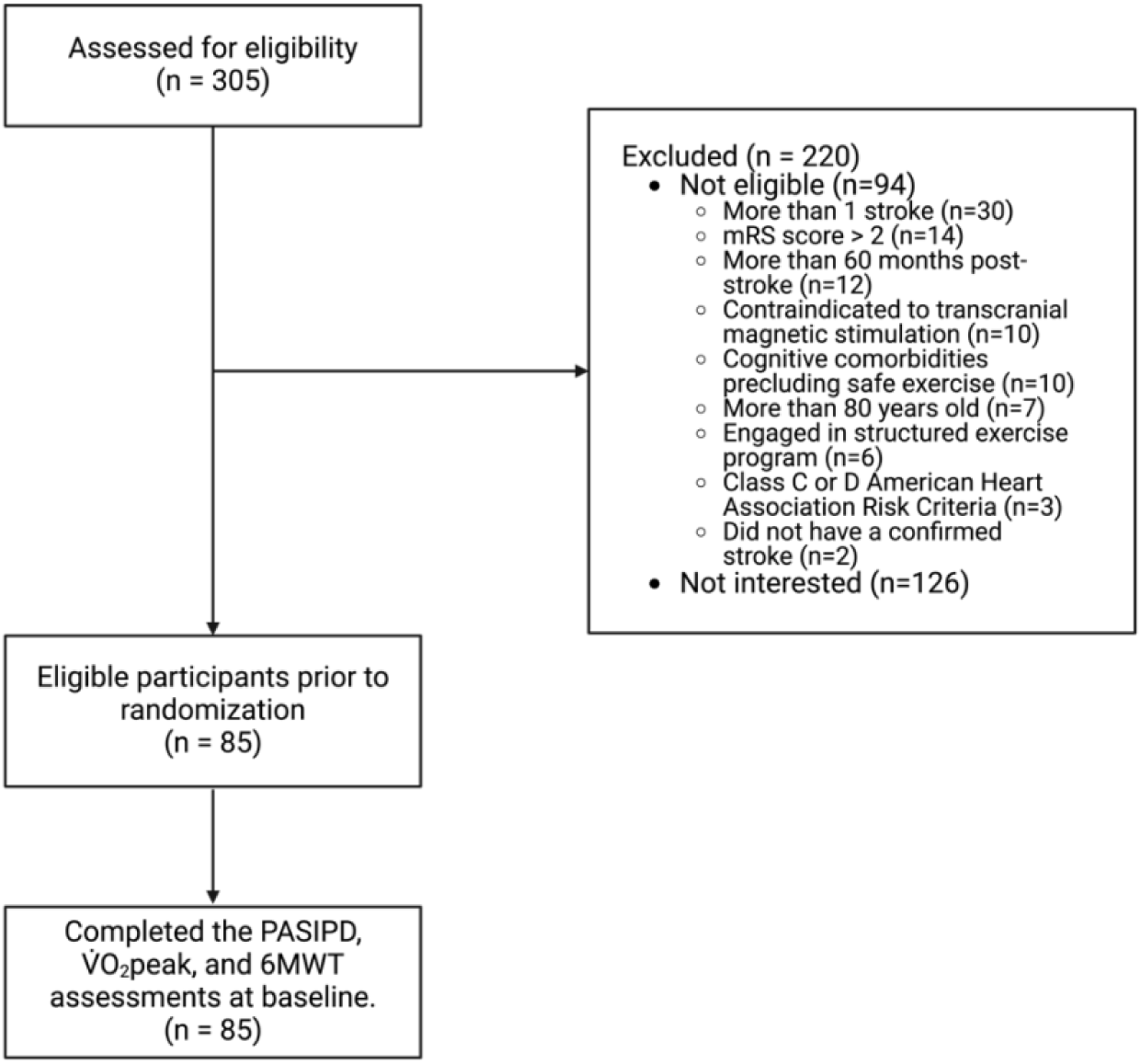
Participant flow chart.

**Table 1.** Demographic variables of all participants and values disaggregated by sex.

| Variable | All | Males | Females |
| --- | --- | --- | --- |
| N | 85 | 53 | 32 |
| Age, years | 65.1 (9.5) | 66.8 (9.4) | 62.2 (9.1)* |
| Height, m | 1.67 (0.09) | 1.71 (0.08) | 1.61 (0.08)* |
| Weight, kg | 80.2 (18.2) | 81.5 (14.7) | 77.8 (22.8) |
| BMI, kg/m<sup>2</sup> | 28.5 (6.7) | 27.6 (4.1) | 30.0 (9.6) |
| Time post-stroke, years | 1.8 (1.2) | 1.6 (1.1) | 2.0 (1.4) |
| Type of stroke, n (%) |  |  |  |
| Ischemic | 59 (69%) | 38 (71%) | 21 (66%) |
| Hemorrhagic | 18 (21%) | 12 (22%) | 6 (18%) |
| Unknown | 8 (10%) | 3 (7%) | 5 (16%) |
| Stroke location, n (%) |  |  |  |
| Cortical | 19 (22%) | 10 (19%) | 9 (28%) |
| Subcortical | 44 (52%) | 29 (55%) | 15 (47%) |
| Cerebellar | 6 (7%) | 4 (8%) | 2 (6%) |
| Brainstem | 9 (11%) | 5 (9%) | 4 (13%) |
| Unknown | 7 (8%) | 5 (9%) | 2 (6%) |
| NIHSS | 2 [3] | 1 [2] | 2 [3] |
| MoCA | 25 [7] | 25 [6] | 27 [7] |
| mRS | 1 [1] | 1 [1] | 1 [1] |
| $\dot{V}O_{2peak}$, mL/kg/min | 17.2 (5.8) | 18.3 (5.7) | 15.3 (5.8)* |
| $\leq 15$ mL/kg/min $\dot{V}O_{2peak}$, n(%) | 35 (42%) | 17 (32%) | 18 (56%)* |
| 6MWT, m | 355.2 (141.9) | 382.6 (141.4) | 309.6 (132.8)* |
| 10-meter usual gait speed, m/s | 1.05 (0.38) | 1.11 (0.37) | 0.96 (0.38) |
| $\leq 350$ m 6MWT, n(%) | 38 (45%) | 18 (34%) | 20 (63%)* |
| Physical activity, MET hours/week | 12.9 (12.3) | 15.3 (13.5) | 9.2 (8.9)* |

Variables are listed as means (standard deviation) or median [interquartile range] unless otherwise reported. \* $p < 0.05$ between sexes. Abbreviations: 6MWT: 6-minute walk test; BMI: body mass index; MET: metabolic equivalent; MoCA: Montreal Cognitive Assessment; NIHSS: National Institute of Health Stroke Severity; $\dot{V}O_{2peak}$ : peak oxygen uptake.

Participants were on average 1.8 (standard deviation [SD] 1.2, range: 0.4 to 4.8) years post-stroke and had mild to moderate stroke severity as assessed by the NIHSS (median = 2, IQR: 3) and modified Rankin Scale (median = 1, IQR: 1). Males were older (mean difference [MD] = 4.6 years, 95% CI [0.4, 8.6], *p*=0.03) and taller (MD = 0.10 m, 95% CI [0.06, 0.14], p<0.001) than females, and had greater V̇O_2_peak (MD = 3.0 mL/kg/min, 95% CI [0.4, 5.6], p=0.02) and 6MWT distance (MD = 73.0 m, 95% CI [11.4, 134.5], p=0.02). In total, 35 participants (42%) were identified as having a V̇O_2_peak <15 mL/kg/min and 38 (45%) participants were identified as having a 6MWT < 350m. Females were more likely to have a V̇O_2_peak < 15 mL/kg/min (p = 0.03) and a 6MWT < 350m (p = 0.01) compared to males. Males also self-reported a higher PA level (MD = 6.1 MET hours/week, 95% [1.2, 10.9], p=0.01). No other significant differences, including differences between stroke type (χ^2^ = 0.26) and location (χ^2^ = 0.83), were observed between the sexes. All participants underwent the clinical assessments over a 2-day period and there were no indeterminate or missing index test or reference standard results. No adverse events occurred during the clinical assessments.

Table 2 presents multivariable logistic regression analyses for V̇O_2_peak (Models 1, 2, 3) adjusted for covariates of age, sex, and 10m usual gait speed. Four outlying covariate patterns were identified in the primary model (Model 1) and subsequently removed given their relatively high influence on the beta-coefficients (∼12% to ∼80%) and model fit (52%). There was no evidence of collinearity in the multivariate model (mean VIF = 1.16). There was a significant association between the V̇O_2_peak ≥15 mL/kg/min cut point and the PASIPD after accounting for age, sex and 10m usual gait speed, explaining 46% of the variance (n=81 observations, *R^2^* = 0.46, *p <* 0.0001). In the adjusted logistic regression model, each one-unit increase in the PASIPD was associated with a 21% increase in the odds of having a V̇O_2_peak ≥ 15 mL/kg/min [aOR (SE) = 1.21 (0.07); 95% CI 1.07, 1.36, p = 0.002]. Sensitivity analyses using a V̇O_2_peak cut point of 12 mL/kg/min demonstrated consistent results, but not a V̇O_2_peak cut point of 18 mL/kg/min (Table 2, Models 2 and 3).

**Table 2.**
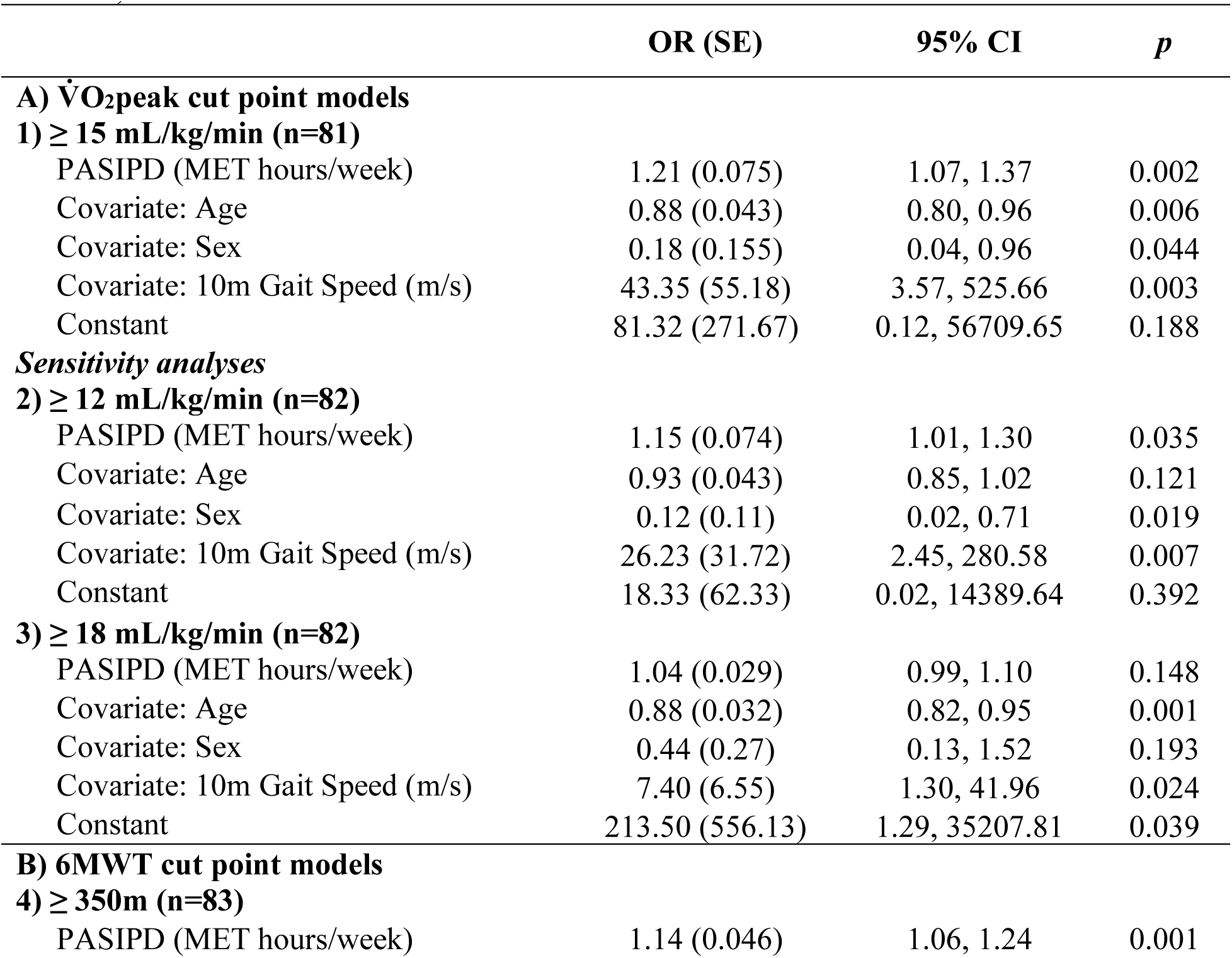
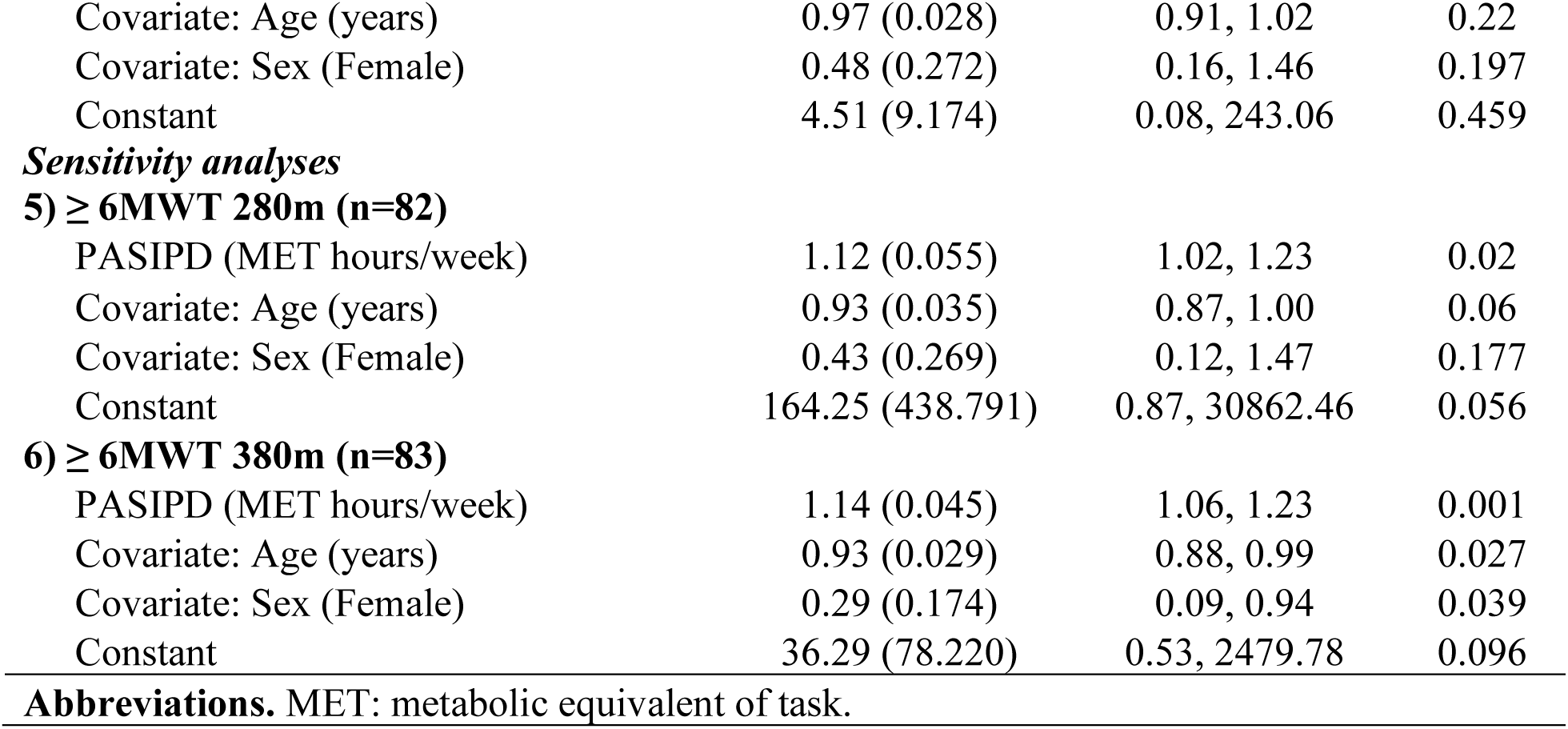
Multivariable logistic regression analyses examining the relationship between the PASIPD (independent continuous variable) and A) cardiorespiratory fitness (V̇O_2_peak, dependent dichotomous variable) and B) 6-minute walk test (6MWT, dependent dichotomous variable).

In the multivariate logistic regression model for 6MWT ≥ 350m, adjusted for covariates of age and sex (Table 2, Model 4), two outlying covariate patterns had an influential effect on the beta-coefficient (6 to 21%) and model fit (28%) and thus were removed from the model. There was no evidence of collinearity in the multivariate model (mean VIF = 1.10). There was a significant association between the 6MWT ≥350 m cutoff and the PASIPD after accounting for age and sex, explaining 22% of the variance (n=83 observations, *R^2^* = 0.22, *p <* 0.0001). In the adjusted logistic regression model, each one-unit increase in the PASIPD was associated with a 14% increase in the odds of having a 6MWT ≥ 350 m [aOR (SE) = 1.14 (0.05); 95% CI 1.05, 1.23, p = 0.001]. Sensitivity analyses using a 6MWT cut point of 280 mL/kg/min and 380 mL/kg/min demonstrated consistent results (Table 2, Models 5 and 6). Figure 2 displays the predicted probabilities of identifying an individual with a V̇O_2_peak value of ≥ 15 mL/kg/min (Panel A) and a 6MWT value of ≥ 350 m (Panel B).

**Figure 2.**
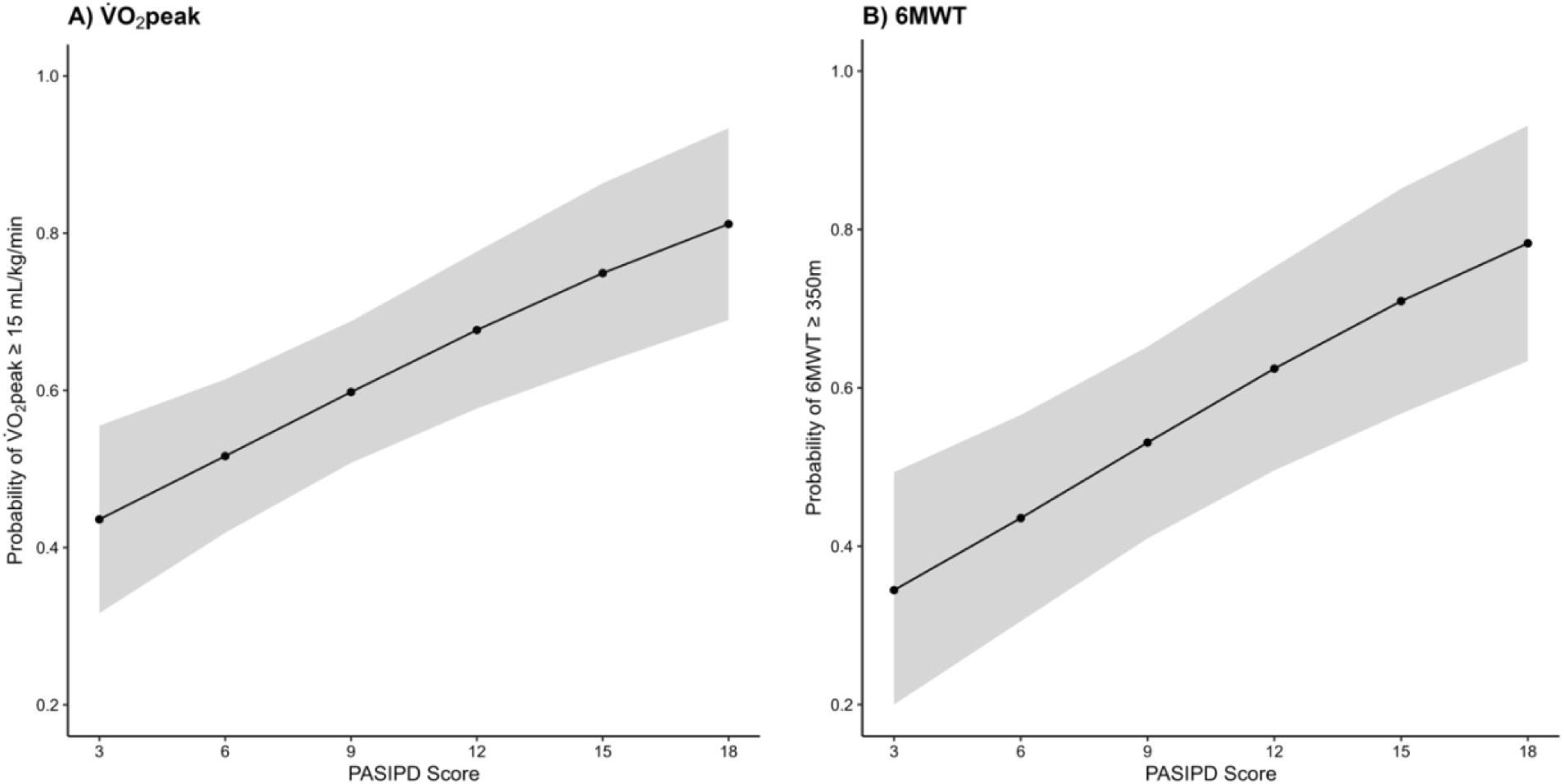
Predicted probabilities of achieving a V̇O_2_peak ≥15 mL/kg/min (Panel A) and 6MWT ≥ 350 m (Panel B) by PASIPD score (MET-hours/week). Shaded area denotes 95% confidence interval bands.

The diagnostic accuracy results of the multivariable logistic regression analyses using a predicted probability cutoff of 0.5 for each V̇O_2_peak and 6MWT cut point are presented in Table 3. Contingency tables for each cut point can be seen in Appendix 2 Tables S1-S6. The primary V̇O_2_peak model (≥ 15 mL/kg/min) adjusted for age, sex and 10-m gait speed had a perfect AUC of 0.91 (95% CI 0.85, 0.97), with a sensitivity of 0.88 (95% CI 0.75, 0.95) and specificity of 0.75 (95% CI 0.57, 0.89), and can correctly classify 83% [0.83 (95% CI 0.73, 0.90)] of individuals with a V̇O_2_peak of ≥ 15 mL/kg/min. The primary 6MWT model (≥ 350 m) adjusted for age and sex, had a perfect AUC of 0.90 (95% CI 0.83, 0.96), with a sensitivity of 0.70 (95% CI 0.55, 0.83) and specificity of 0.67 (95% CI 0.49, 0.81), and can correctly classify 69% [0.69 (95% CI 0.58, 0.78)] of individuals with a 6MWT of ≥ 350 m. Sensitivity analysis for the additional V̇O_2_peak cut points (12 and 18 mL/kg/min) and 6MWT cut points (280 and 380 m) demonstrate excellent to perfect AUC values.

**Table 3.** Diagnostic performance of the adjusted multivariable logistic regression models for each V̇O_2_peak and 6MWT cut point.

| A) $\dot{V}O_{2peak}$ | 1) $\geq 15$ mL/kg/min | 2) $\geq 12$ mL/kg/min | 3) $\geq 18$ mL/kg/min |
| --- | --- | --- | --- |
| Area under the curve | 0.91 (0.85, 0.97) | 0.88 (0.81, 0.95) | 0.82 (0.74, 0.91) |
| Sensitivity | 0.88 (0.75, 0.95) | 0.92 (0.83, 0.97) | 0.60 (0.42, 0.76) |
| Specificity | 0.75 (0.57, 0.89) | 0.33 (0.13, 0.59) | 0.81 (0.67, 0.91) |
| NPV | 0.84 (0.71, 0.93) | 0.83 (0.72, 0.91) | 0.70 (0.51, 0.85) |
| NPV | 0.80 (0.61, 0.92) | 0.55 (0.23, 0.83) | 0.73 (0.59, 0.84) |
| Positive LR | 3.51 (1.91, 6.45) | 1.38 (0.99, 1.93) | 3.13 (1.64, 5.98) |
| Negative LR | 0.16 (0.08, 0.35) | 0.23 (0.08, 0.68) | 0.49 (0.32, 0.76) |
| Correctly classified | 0.83 (0.73, 0.90) | 0.79 (0.69, 0.87) | 0.72 (0.61, 0.81) |
| B) 6MWT | 4) $\geq 350$ m | 5) $\geq 280$ m | 6) $\geq 380$ m |
| Area under the curve | 0.90 (0.83, 0.96) | 0.91 (0.83, 0.98) | 0.93 (0.88, 0.98) |
| Sensitivity | 0.70 (0.55, 0.83) | 0.98 (0.91, 1.00) | 0.64 (0.48, 0.78) |
| Specificity | 0.67 (0.49, 0.81) | 0.35 (0.15, 0.59) | 0.80 (0.65, 0.91) |
| PPV | 0.73 (0.58, 0.85) | 0.82 (0.72, 0.90) | 0.77 (0.60, 0.90) |
| NPV | 0.63 (0.46, 0.78) | 0.88 (0.47, 1.00) | 0.69 (0.54, 0.81) |
| Positive LR | 2.11 (1.28, 3.47) | 1.51 (1.10, 2.09) | 3.29 (1.70, 6.38) |
| Negative LR | 0.45 (0.27, 0.73) | 0.05 (0.01, 0.35) | 0.44 (0.29, 0.68) |
| Correctly classified | 0.69 (0.58, 0.78) | 0.83 (0.73, 0.90) | 0.72 (0.61, 0.82) |

Data are presented as value (95% CI). **Abbreviations.** 6MWT: 6-minute walk test; LR: likelihood ratio; NPV: negative predictive value; PPV: positive predictive value; $\dot{V}O_{2peak}$ : peak oxygen uptake.

Table 4 illustrates the ROC results of the Youden-derived PASIPD cut points for identifying individuals with stroke with V̇O_2_peak fitness (e.g., ≥ 15 mL/kg/min [primary cut point], ≥ 12 mL/kg/min, and ≥ 18 mL/kg/min [sensitivity cut points]) and 6MWT impairments (e.g., ≥ 350 m [primary cut point], ≥ 280 m, and ≥ 380 m [sensitivity cut points]). Contingency tables for each cut point and Youden-derived PASIPD cut point can be seen in Appendix 2 Tables S7-S12. The optimal PASIPD cut point that maximizes both sensitivity and specificity to identify a V̇O_2_peak cut point ≥ 15 mL/kg/min was 8.9 MET hours/week. This cut point demonstrated a borderline acceptable AUC of 0.69 (95% CI 0.59, 0.79) with a sensitivity of 0.77 (95% CI 0.62, 0.89) and specificity of 0.61 (95% CI 0.45, 0.76). For identifying 6MWT distance of ≥ 350 meters, the optimal PASIPD cut point was 10.6 MET hours/week. This specific value demonstrated an acceptable AUC of 0.75 (95% CI: 0.65, 0.86), with a sensitivity of 0.62 (95% CI 0.46, 0.75) and specificity of 0.82 (95% CI 0.66, 0.92). Sensitivity analysis for additional optimal Youden-derived PASIPD cut points can be seen in Table 4. Figure S1 in Appendix 3 displays the ROC curves for the Youden-derived PASIPD cut point for each V̇O_2_peak and 6MWT cut points.

**Table 4.** Receiver operator characteristic and diagnostic parameters of the Youden-derived PASIPD cut points for each V̇O_2_peak and 6MWT cut points.

| A) $\dot{V}O_{2\text{peak}}$ | 1) $\geq 15$ mL/kg/min | 2) $\geq 12$ mL/kg/min | 3) $\geq 18$ mL/kg/min |
| --- | --- | --- | --- |
| PASIPD cut point | 8.9 MET-hrs/wk | 6.1 MET-hrs/wk | 3.6 MET-hrs/wk |
| Area under the curve | 0.69 (0.59, 0.79) | 0.68 (0.56, 0.80) | 0.67 (0.59, 0.75) |
| Sensitivity | 0.77 (0.62, 0.89) | 0.86 (0.74, 0.94) | 0.55 (0.42, 0.67) |
| Specificity | 0.61 (0.45, 0.76) | 0.45 (0.26, 0.64) | 0.90 (0.70, 0.99) |
| PPV | 0.68 (0.53, 0.80) | 0.75 (0.63, 0.85) | 0.95 (0.82, 0.99) |
| NPV | 0.71 (0.54, 0.85) | 0.62 (0.38, 0.82) | 0.40 (0.26, 0.55) |
| Positive LR | 1.98 (1.31, 3.00) | 1.55 (1.10, 2.19) | 5.74 (1.51, 21.86) |
| Negative LR | 0.37 (0.21, 0.68) | 1.55 (1.10, 2.19) | 0.50 (0.37, 0.68) |
| Correctly classified | 0.69 (0.58, 0.79) | 0.72 (0.61, 0.81) | 0.64 (0.52, 0.74) |
| B) 6MWT | 4) $\geq 350$ m | 5) $\geq 280$ m | 6) $\geq 380$ m |
| PASIPD cut point | 10.6 MET-hrs/wk | 6.5 MET-hrs/wk | 10.6 MET-hrs/wk |
| Area under the curve | 0.71 (0.62, 0.81) | 0.68 (0.56, 0.79) | 0.74 (0.64, 0.83) |
| Sensitivity | 0.81 (0.64, 0.92) | 0.85 (0.72, 0.93) | 0.78 (0.61, 0.90) |
| Specificity | 0.63 (0.48, 0.77) | 0.45 (0.28, 0.64) | 0.71 (0.57, 0.83) |
| PPV | 0.62 (0.46, 0.75) | 0.71 (0.58, 0.82) | 0.67 (0.50, 0.80) |
| NPV | 0.82 (0.66, 0.92) | 0.65 (0.43, 0.84) | 0.81 (0.67, 0.92) |
| Positive LR | 2.19 (1.47, 3.27) | 1.55 (1.11, 2.16) | 2.72 (1.69, 4.38) |
| Negative LR | 0.31 (0.15, 0.62) | 0.34 (0.16, 0.71) | 0.31 (0.16, 0.59) |
| Correctly classified | 0.71 (0.60, 0.80) | 0.69 (0.58, 0.79) | 0.74 (0.63, 0.83) |

Data are presented as value (95% CI). **Note:** these values are unadjusted values.
**Abbreviations.** 6MWT: 6-minute walk test; LR: likelihood ratio; MET-hrs/week: metabolic equivalents of task hours per week; NPV: negative predictive value; PPV: positive predictive value; $\dot{V}O_{2\text{peak}}$ : peak oxygen uptake.

## Discussion

This study demonstrated that self-reported PA measured using the PASIPD was positively and significantly associated with gold-standard assessment of V̇O_2_peak in the chronic phase of stroke recovery (Figure 2, Panel A). Each one-unit increase in the PASIPD was associated with a 21% increase in the odds of having a V̇O_2_peak ≥ 15 mL/kg/min [Model 1, aOR = 1.21; 95% CI 1.07, 1.36, p = 0.002]. Our sensitivity analysis suggest that each one-unit increase in the PASIPD (1 MET-hours/week) was associated with a 15% increase in the odds of having a V̇O_2_peak ≥ 12 mL/kg/min [Model 2, aOR = 1.15; 95% CI 1.01, 1.30, p = 0.035], but no significant relationship was found for a V̇O_2_peak cut point of ≥ 18 mL/kg/min [Model 3, aOR = 1.04; 95% CI 0.99, 1.10, p = 0.148]. This suggests that the PASIPD may only have clinical utility in identifying individuals who are more likely to have lower V̇O_2_peak values (12 to 15 mL/kg/min). Nonetheless, our findings confirm our primary hypothesis and extends prior research which found fair to modest relationships between V̇O_2_peak and PA.^16^

Interestingly, we also found positive and significant associations between the PASIPD and walking capacity, which refutes our evidence-based secondary hypothesis^17^ (Figure 2, Panel B). Each one-unit increase in the PASIPD (1 MET-hours/week) was associated with a 14% increase in the odds of having a 6MWT ≥ 350 mL/kg/min [Model 4, aOR (SE) = 1.14 (0.05); 95% CI 1.05, 1.23, p = 0.001], which was consistent with sensitivity analyses using 6MWT cut point of 280 mL/kg/min and 380 mL/kg/min (12-14% increase odds). Indeed, walking capacity is considered an important activity limitation that is related to objective PA,^9,16^ and our study contrasts prior literature^17^ and provides new evidence to support that self-reported PA is associated with walking capacity in the chronic phase of stroke recovery, at least in individuals with mild to moderate levels of disability. Future research is warranted to evaluate the relationship between self-reported PA and other field tests, such as the incremental shuttle walk test, as individuals post-stroke report that deficits in walking distance is a key factor for limiting engagement in community activities.^43^

Using a predicted probability cutoff of 0.5 in our multivariable logistic regression models, we also found that the V̇O_2_peak (12-18 mL/kg/min) and 6MWT (≥280-380 m) cut points demonstrate excellent to perfect diagnostic accuracy (AUC: 0.82-0.93, Table 3). These results indicate that the models accurately identify participants with adequate V̇O_2_peak (72-83% correctly classified) and walking capacity (69-83% correctly classified). To facilitate clinical implementation of the PASIPD, we identified optimal PASIPD cut points (MET-hours/week) via ROC analyses that balance sensitivity, specificity, positive predictive value and positive likelihood ratio (Table 4). In contrast to our multivariable logistic models, these cut points do not account for confounders (e.g., age, sex, 10m gait speed) and thus should be interpreted with caution. Nonetheless, with overall moderate diagnostic accuracy, a PASIPD cut point of 6.1-8.9 MET-hours/week may be used to discriminate individuals post-stroke who have a V̇O_2_peak 12-15 mL/kg/min, while a cut point between 6.5-10.6 MET-hours/week discriminates individuals with a 6MWT of 280-380m.

The findings of this study have practical implications for stroke clinical practice. Indeed, stroke physiotherapists have reported barriers with implementing AE^12,13^ and require tools that can assist with AE screening.^44,45^ Our findings provide psychometric evidence that the PASIPD may be a useful self-reported tool that can be used by clinicians in identifying individuals with varying levels of V̇O_2_peak and 6MWT impairments post-stroke. Integrating the 13-item self-reported PASIPD questionnaire^14^ may provide clinicians with an accurate, inexpensive, and efficient tool that may be used during assessments to help guide clinical decision making or to identify individuals who may need further in-depth CPET or targeted intervention. Nevertheless, clinicians are encouraged to continue utilizing self-reported assessments with objective assessment where feasible^11^ and to continue adopting best practice guidelines for exercise.^7,8,46^ Future research is required to validate these cut points in other stroke rehabilitation contexts and to evaluate the feasibility of implementing our PASIPD cut points in stroke clinical practice.

It is also important to recognize that despite our participants having mild stroke severity and above-average gait speeds (1.05 m/s vs. 0.84 m/s)^47^ and walking capacities (6MWT 359.1 ± 141.1 vs 284 ± 107 m),^48^ a significant proportion of our participants were deemed to have low self-reported PA levels (12.9 ± 12.3 MET-hours/week) and low V̇O_2_peak as determined by our evidence-based cut point of 15 mL/kg/min (n=35/85, 42%). Given the prognostic importance of V̇O_2_peak for stroke recurrence and all-cause mortality,^1–3^ our findings highlight the need for appropriate screening tools that can be used to identify individuals at risk for low V̇O_2_peak or those in need of personalized exercise support.

There are important limitations that need to be considered to interpret the findings of our study. First, we used a 13-item self-reported PASIPD questionnaire that relies on information regarding leisure time activity, household and work-related activity and exercise behaviours over the previous 7 days, which could result in recall bias of PA. However, the PASIPD has good test-retest reliability^15^ and criterion validity that is comparable to well established self-report PA questionnaires from the general population.^49^ Wearable technologies are a fast and emerging field of research, but are costly to implement in clinical practice, thus the use of simple and brief questionnaires can offer practical solutions to measure PA after stroke. However, future research is warranted to examine accelerometer derived PA cut points and their relationship with V̇O_2_peak and 6MWT impairments after stroke.

## Conclusions

In conclusion, we found that self-reported PA, as measured by the PASIPD, was positively and significantly associated with V̇O_2_peak and 6MWT among individuals post-stroke. We also identified optimal PASIPD cut points that clinicians may use to identify individuals post-stroke who may be at risk of having low V̇O_2_peak and 6MWT impairments, and those who may require more in-depth cardiopulmonary exercise testing.

## Data Availability

All data produced in the present study are available upon reasonable request to the authors.

## Acknowledgements

The authors would like to acknowledge Hanna Fang for her assistance with study tasks, recruitment, data collection, and exercise training.

## Disclosures & Sources of funding received

KM is supported by CANTRAIN Clinical Trials Fellowship. LR is supported by a Stroke Cog Postdoctoral Fellowship. EW is supported by a Michael Smith Health Research BC Trainee Award and a StrokeCog Postdoctoral Fellowship. KSN is supported by a Mitacs Accelerate Postdoctoral Internship. JJE is supported by the Canada Research Chairs program. MR is supported by a Salary Award (Junior II) from Fonds de Recherche Santé Québec. This study is funded by an operating grant from the Canadian Institutes of Health Research (388320). AT reports grants from Canadian Institutes of Health Research; grants from Physiotherapy Foundation of Canada; and grants from Heart and Stroke Foundation of Canada

## Supplemental appendices

## Appendix 1 STARD Checklist

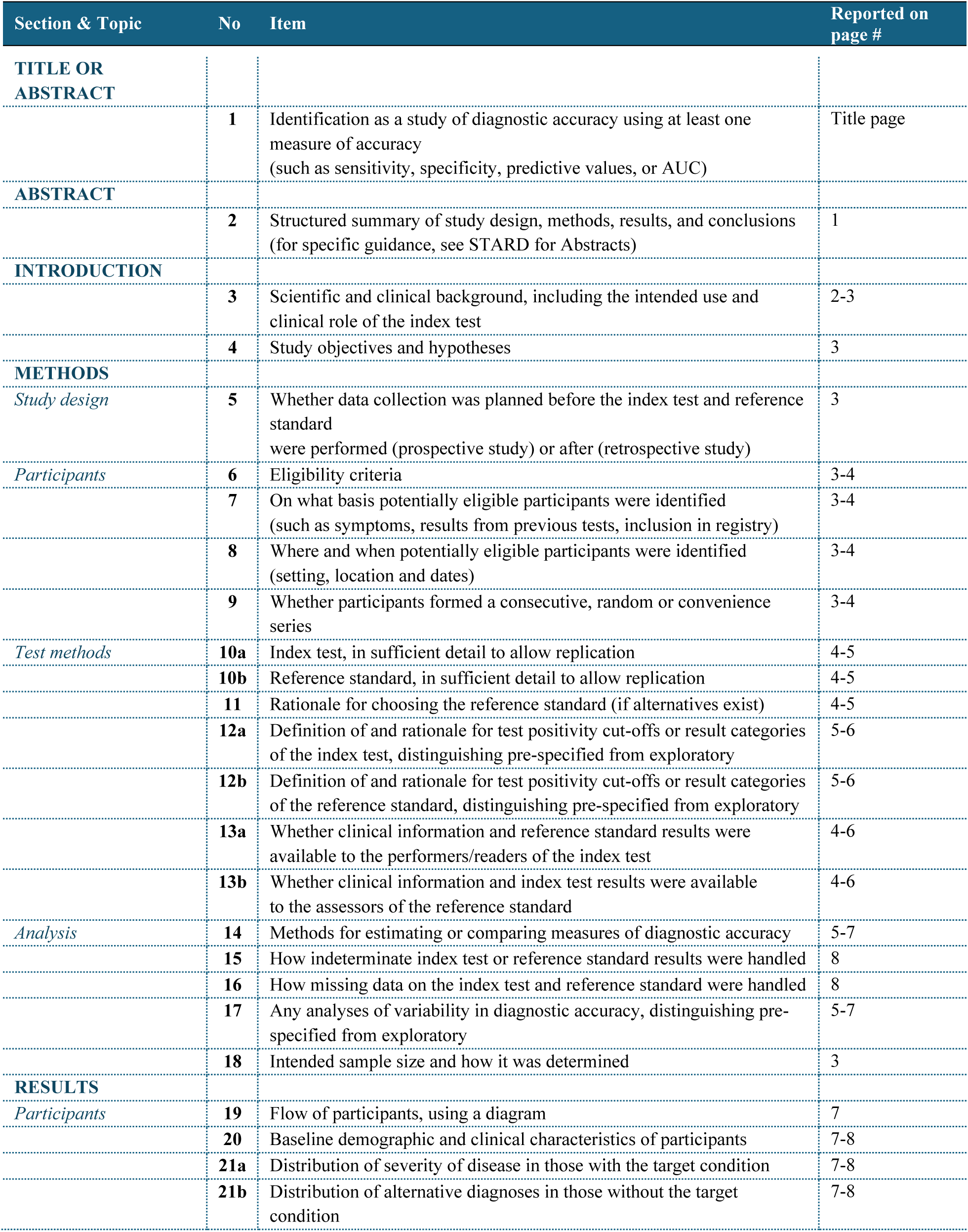
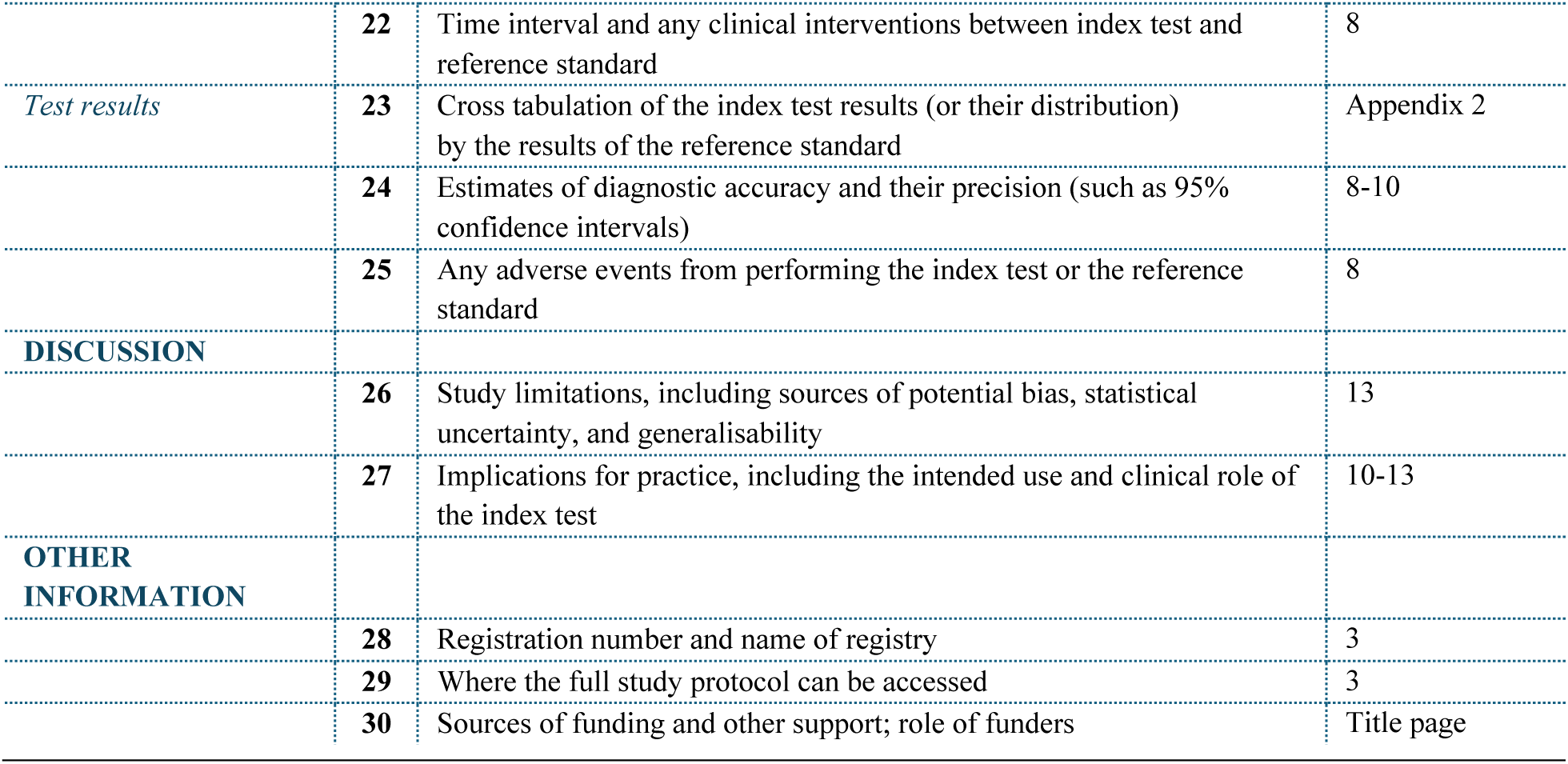

Appendix 1 – STARD Checklist
| Section & Topic | No | Item | Reported on page # |
| --- | --- | --- | --- |
| **TITLE OR ABSTRACT** |  |  |  |
|  | **1** | Identification as a study of diagnostic accuracy using at least one measure of accuracy (such as sensitivity, specificity, predictive values, or AUC) | Title page |
| **ABSTRACT** |  |  |  |
|  | **2** | Structured summary of study design, methods, results, and conclusions (for specific guidance, see STARD for Abstracts) | 1 |
| **INTRODUCTION** |  |  |  |
|  | **3** | Scientific and clinical background, including the intended use and clinical role of the index test | 2-3 |
|  | **4** | Study objectives and hypotheses | 3 |
| **METHODS** |  |  |  |
| *Study design* | **5** | Whether data collection was planned before the index test and reference standard were performed (prospective study) or after (retrospective study) | 3 |
| *Participants* | **6** | Eligibility criteria | 3-4 |
|  | **7** | On what basis potentially eligible participants were identified (such as symptoms, results from previous tests, inclusion in registry) | 3-4 |
|  | **8** | Where and when potentially eligible participants were identified (setting, location and dates) | 3-4 |
|  | **9** | Whether participants formed a consecutive, random or convenience series | 3-4 |
| *Test methods* | **10a** | Index test, in sufficient detail to allow replication | 4-5 |
|  | **10b** | Reference standard, in sufficient detail to allow replication | 4-5 |
|  | **11** | Rationale for choosing the reference standard (if alternatives exist) | 4-5 |
|  | **12a** | Definition of and rationale for test positivity cut-offs or result categories of the index test, distinguishing pre-specified from exploratory | 5-6 |
|  | **12b** | Definition of and rationale for test positivity cut-offs or result categories of the reference standard, distinguishing pre-specified from exploratory | 5-6 |
|  | **13a** | Whether clinical information and reference standard results were available to the performers/readers of the index test | 4-6 |
|  | **13b** | Whether clinical information and index test results were available to the assessors of the reference standard | 4-6 |
| *Analysis* | **14** | Methods for estimating or comparing measures of diagnostic accuracy | 5-7 |
|  | **15** | How indeterminate index test or reference standard results were handled | 8 |
|  | **16** | How missing data on the index test and reference standard were handled | 8 |
|  | **17** | Any analyses of variability in diagnostic accuracy, distinguishing pre-specified from exploratory | 5-7 |
|  | **18** | Intended sample size and how it was determined | 3 |
| **RESULTS** |  |  |  |
| *Participants* | **19** | Flow of participants, using a diagram | 7 |
|  | **20** | Baseline demographic and clinical characteristics of participants | 7-8 |
|  | **21a** | Distribution of severity of disease in those with the target condition | 7-8 |
|  | **21b** | Distribution of alternative diagnoses in those without the target condition | 7-8 |
|  | 22 | Time interval and any clinical interventions between index test and reference standard | 8 |
| *Test results* | 23 | Cross tabulation of the index test results (or their distribution) by the results of the reference standard | Appendix 2 |
|  | 24 | Estimates of diagnostic accuracy and their precision (such as 95% confidence intervals) | 8-10 |
|  | 25 | Any adverse events from performing the index test or the reference standard | 8 |
| **DISCUSSION** |  |  |  |
|  | 26 | Study limitations, including sources of potential bias, statistical uncertainty, and generalisability | 13 |
|  | 27 | Implications for practice, including the intended use and clinical role of the index test | 10-13 |
| **OTHER INFORMATION** |  |  |  |
|  | 28 | Registration number and name of registry | 3 |
|  | 29 | Where the full study protocol can be accessed | 3 |
|  | 30 | Sources of funding and other support; role of funders | Title page |

## Appendix 2 Contigency Tables

**Table S1.** Contigency table for V̇O2peak Classification (≥15 mL/kg/min) Using the Adjusted Logistic Regression Model.

Appendix 2 – Contingency Tables
| Table S1. Contingency table for $\dot{V}O_2$peak Classification ($\geq 15$ mL/kg/min) Using the Adjusted Logistic Regression Model | | | |
| --- | --- | --- | --- |
|  | True |  |  |
| Classified | Positive<br>($\dot{V}O_2$peak $\geq 15$ mL/kg/min) | Negative<br>($\dot{V}O_2$peak $< 15$ mL/kg/min) | Total |
| Positive | 43 | 8 | 51 |
| Negative | 6 | 24 | 30 |
| Total | 49 | 32 | 81 |
| Note: Predicted classifications were determined using a probability cutoff of 0.5, such that participants with a predicted probability $\geq 0.5$ were classified as having a $\dot{V}O_2$peak $\geq 15$ mL/kg/min. The true $\dot{V}O_2$peak status was defined based on measured values whereby individuals with a $\dot{V}O_2$peak $\geq 15$ mL/kg/min were considered to have adequate cardiorespiratory fitness (positive), whereas those with a $\dot{V}O_2$peak $< 15$ mL/kg/min were considered to have reduced cardiorespiratory fitness (negative). | | | |

**Table S2.** Contigency table for V̇O_2_peak Classification (≥12 mL/kg/min) Using the Adjusted Logistic Regression Model.

| Table S2. Contingency table for $\dot{V}O_2$peak Classification ($\geq 12$ mL/kg/min) Using the Adjusted Logistic Regression Model | | | |
| --- | --- | --- | --- |
|  | True |  |  |
| Classified | Positive<br>($\dot{V}O_2$peak $\geq 12$ mL/kg/min) | Negative<br>($\dot{V}O_2$peak $< 12$ mL/kg/min) | Total |
| Positive | 59 | 12 | 71 |
| Negative | 5 | 6 | 11 |
| Total | 64 | 18 | 82 |

Note: Predicted classifications were determined using a probability cutoff of 0.5, such that participants with a predicted probability $\geq 0.5$ were classified as having a $\dot{V}O_{2peak} \geq 12$ mL/kg/min. The true $\dot{V}O_{2peak}$ status was defined based on measured values whereby individuals with a $\dot{V}O_{2peak} \geq 12$ mL/kg/min were considered to have adequate cardiorespiratory fitness (positive), whereas those with a $\dot{V}O_{2peak} < 12$ mL/kg/min were considered to have reduced cardiorespiratory fitness (negative).

**Table S3.** Contigency table for V̇O_2_peak Classification (≥18 mL/kg/min) Using the Adjusted Logistic Regression Model.

| Classified | True |  | Total |
| --- | --- | --- | --- |
| | Positive<br>($\dot{V}O_{2peak} \geq 18$ mL/kg/min) | Negative<br>($\dot{V}O_{2peak} < 18$ mL/kg/min) | |
| Positive | 21 | 9 | 30 |
| Negative | 14 | 38 | 52 |
| Total | 35 | 47 | 82 |

Note: Predicted classifications were determined using a probability cutoff of 0.5, such that participants with a predicted probability $\geq 0.5$ were classified as having a $\dot{V}O_{2peak} \geq 18$ mL/kg/min. The true $\dot{V}O_{2peak}$ status was defined based on measured values whereby individuals with a $\dot{V}O_{2peak} \geq 18$ mL/kg/min were considered to have adequate cardiorespiratory fitness (positive), whereas those with a $\dot{V}O_{2peak} < 18$ mL/kg/min were considered to have reduced cardiorespiratory fitness (negative).

**Table S4.** Contigency table for 6MWT Classification (≥ 350 m) Using the Adjusted Logistic Regression Model.

| Classified | True |  | Total |
| --- | --- | --- | --- |
| | Positive<br>(6MWT $\geq 350$ m) | Negative<br>(6MWT $< 350$ m) | |
| Positive | 33 | 12 | 45 |
| Negative | 14 | 24 | 38 |
| Total | 47 | 36 | 83 |

Note: Predicted classifications were determined using a probability cutoff of 0.5, such that participants with a predicted probability $\geq 0.5$ were classified as having a 6MWT $\geq 350$ m. The true 6MWT status was defined based on measured values whereby individuals with a 6MWT $\geq 350$ m were considered to have adequate walking capacity (positive), whereas those with a 6MWT $< 350$ m were considered to have reduced walking capacity (negative).

**Table S5.** Contigency table for 6MWT Classification (≥ 280 m) Using the Adjusted Logistic Regression Model.

| Classified | True |  | Total |
| --- | --- | --- | --- |
| | Positive<br>(6MWT $\geq 280$ m) | Negative<br>(6MWT $< 280$ m) | |
| Positive | 61 | 13 | 74 |
| Negative | 1 | 7 | 8 |
| Total | 62 | 20 | 82 |

Note: Predicted classifications were determined using a probability cutoff of 0.5, such that participants with a predicted probability $\geq 0.5$ were classified as having a 6MWT $\geq 280$ m. The true 6MWT status was defined based on measured values whereby individuals with a 6MWT $\geq 280$ m were considered to have adequate walking capacity (positive), whereas those with a 6MWT $< 280$ m were considered to have reduced walking capacity (negative).

**Table S6.**
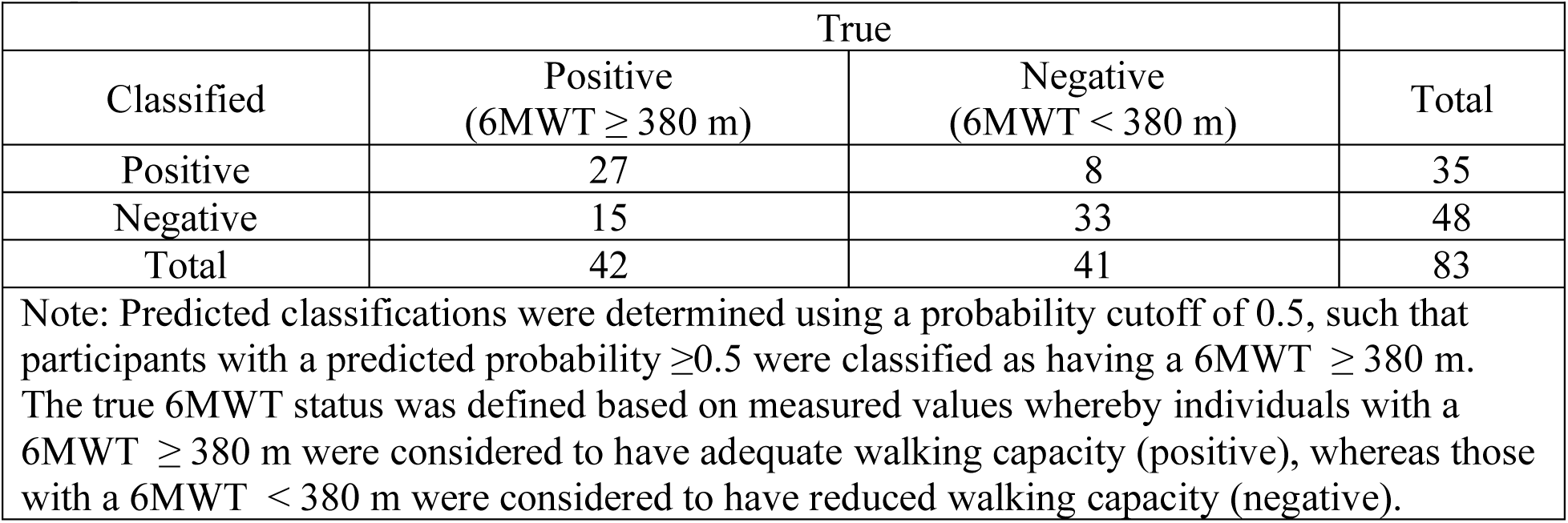
Contigency table for 6MWT Classification (≥ 380 m) Using the Adjusted Logistic Regression Model.

| Classified | True |  | Total |
| --- | --- | --- | --- |
| | Positive<br>(6MWT $\geq 380$ m) | Negative<br>(6MWT $< 380$ m) | |
| Positive | 27 | 8 | 35 |
| Negative | 15 | 33 | 48 |
| Total | 42 | 41 | 83 |

Note: Predicted classifications were determined using a probability cutoff of 0.5, such that participants with a predicted probability $\geq 0.5$ were classified as having a 6MWT $\geq 380$ m. The true 6MWT status was defined based on measured values whereby individuals with a 6MWT $\geq 380$ m were considered to have adequate walking capacity (positive), whereas those with a 6MWT $< 380$ m were considered to have reduced walking capacity (negative).

**Table S7.**
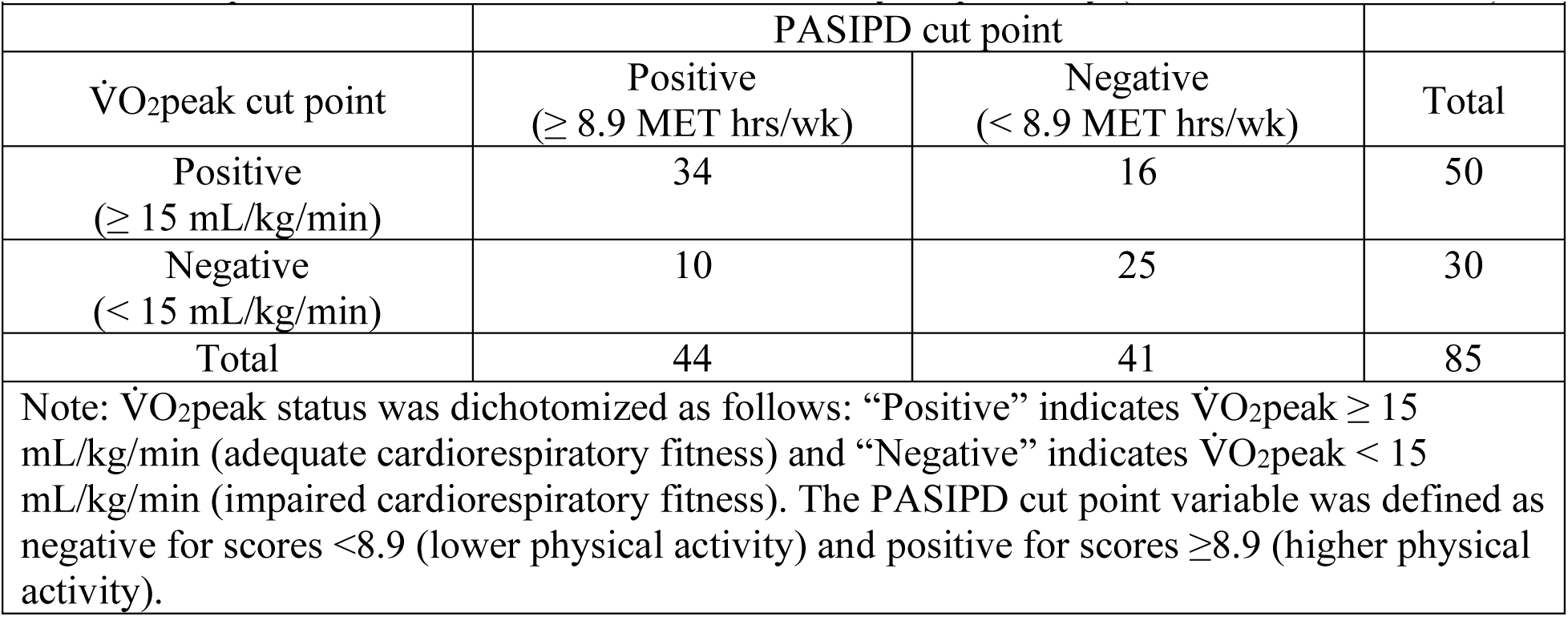
Contigency table for V̇O_2_peak Classification (≥15 mL/kg/min) and Youden-derived PASIPD cut point that maximizes the sum of sensitivy 1-specificity (≥8.9 MET hours/week)

**Table S8.**
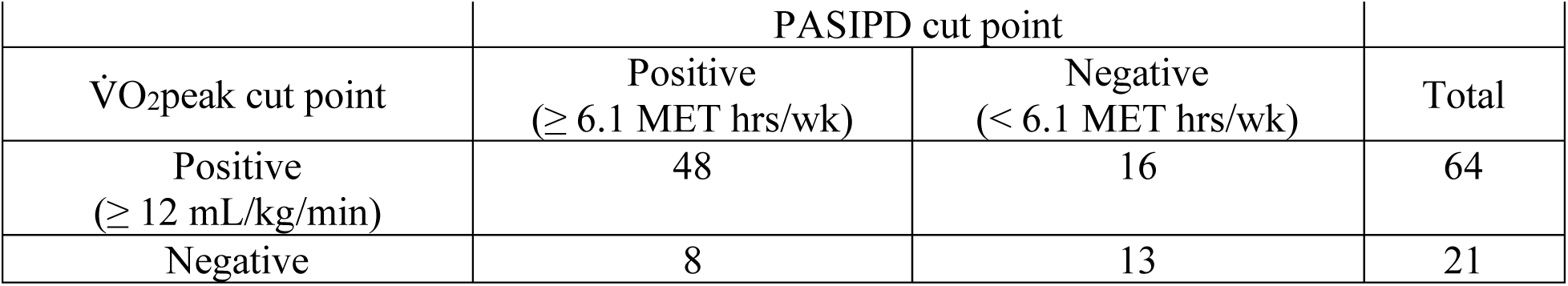
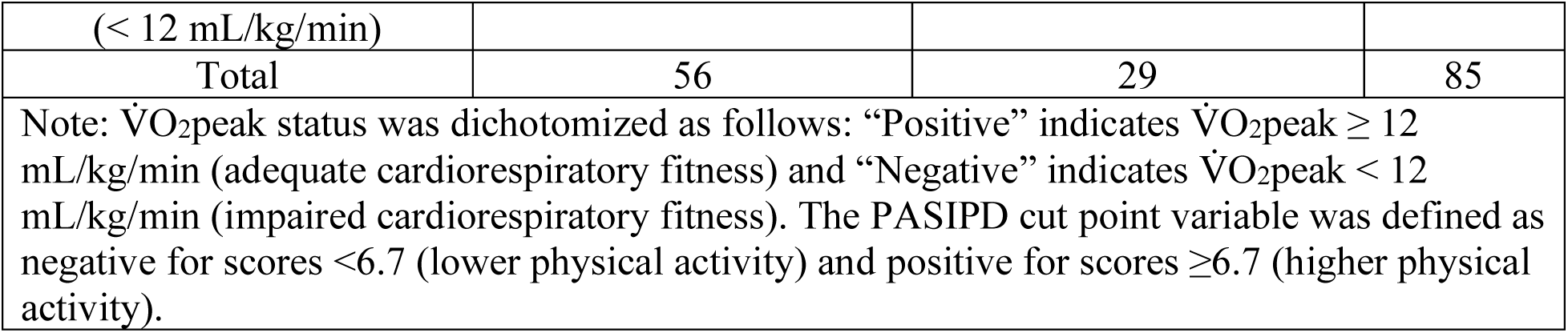
Contigency table for V̇O_2_peak Classification (≥12 mL/kg/min) and Youden-derived PASIPD cut point that maximizes the sum of sensitivy 1-specificity (≥ 6.1 MET hours/week)

**Table S9.** Contigency table for V̇O_2_peak Classification (≥18 mL/kg/min) and Youden-derived PASIPD cut point that maximizes the sum of sensitivy 1-specificity (≥ 3.6 MET hours/week)

| Table S9. Contingency table for $\dot{V}O_{2peak}$ Classification ($\geq 18$ mL/kg/min) and Youden-derived PASIPD cut point that maximizes the sum of sensitivity 1-specificity ($\geq 3.6$ MET hours/week) | | | |
| --- | --- | --- | --- |
|  | PASIPD cut point |  |  |
| $\dot{V}O_{2peak}$ cut point | Positive<br>($\geq 3.6$ MET hrs/wk) | Negative<br>($< 3.6$ MET hrs/wk) | Total |
| Positive<br>($\geq 18$ mL/kg/min) | 35 | 2 | 37 |
| Negative<br>($< 18$ mL/kg/min) | 29 | 19 | 48 |
| Total | 64 | 21 | 85 |
| Note: $\dot{V}O_{2peak}$ status was dichotomized as follows: “Positive” indicates $\dot{V}O_{2peak} \geq 18$ mL/kg/min (adequate cardiorespiratory fitness) and “Negative” indicates $\dot{V}O_{2peak} < 18$ mL/kg/min (impaired cardiorespiratory fitness). The PASIPD cut point variable was defined as negative for scores <3.6 (lower physical activity) and positive for scores $\geq 3.6$ (higher physical activity). | | | |

**Table S10.**
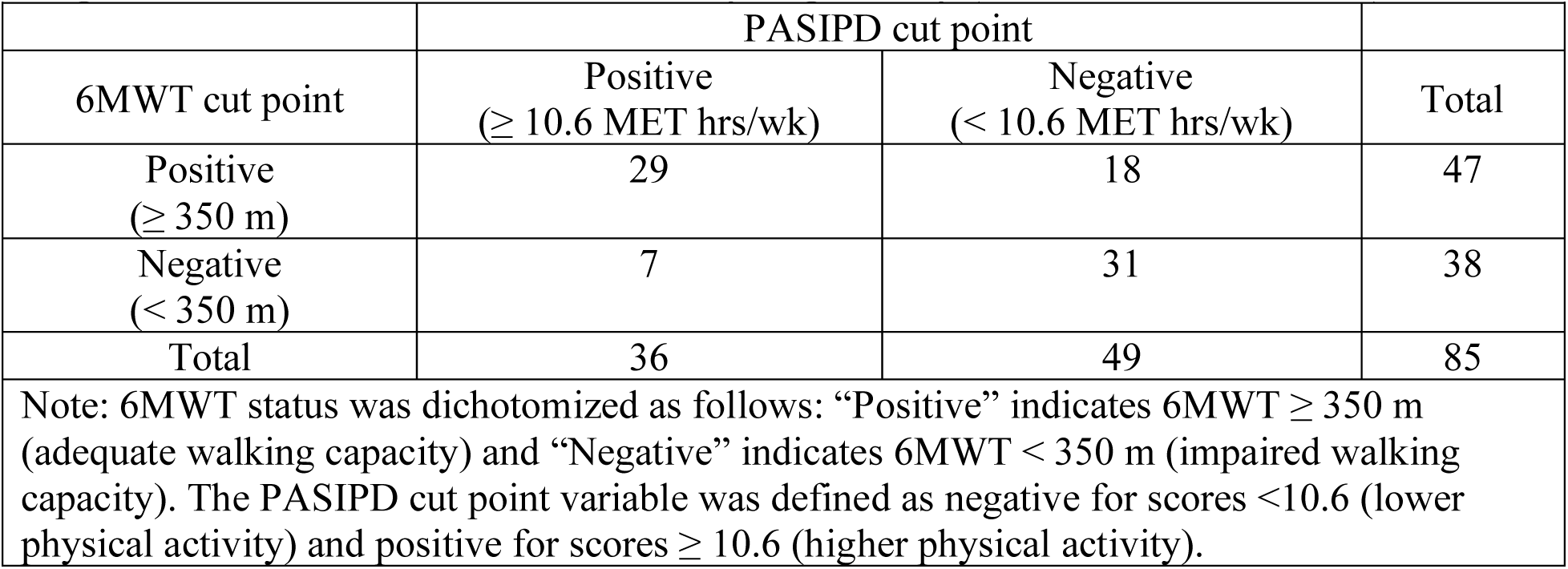
Contigency table for 6MWT Classification (≥350 m) and Youden-derived PASIPD cut point that maximizes the sum of sensitivy 1-specificity (≥ 10.6 MET hours/week)

**Table S11.**
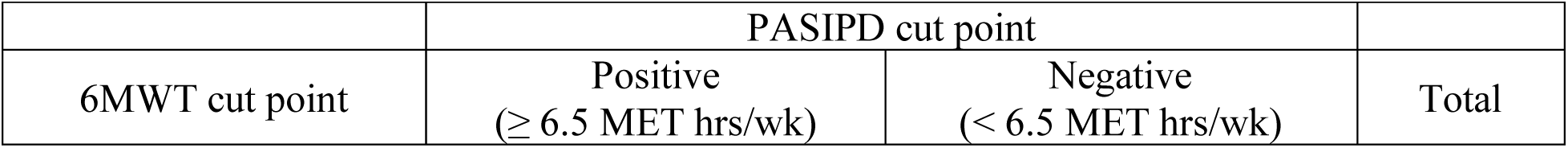
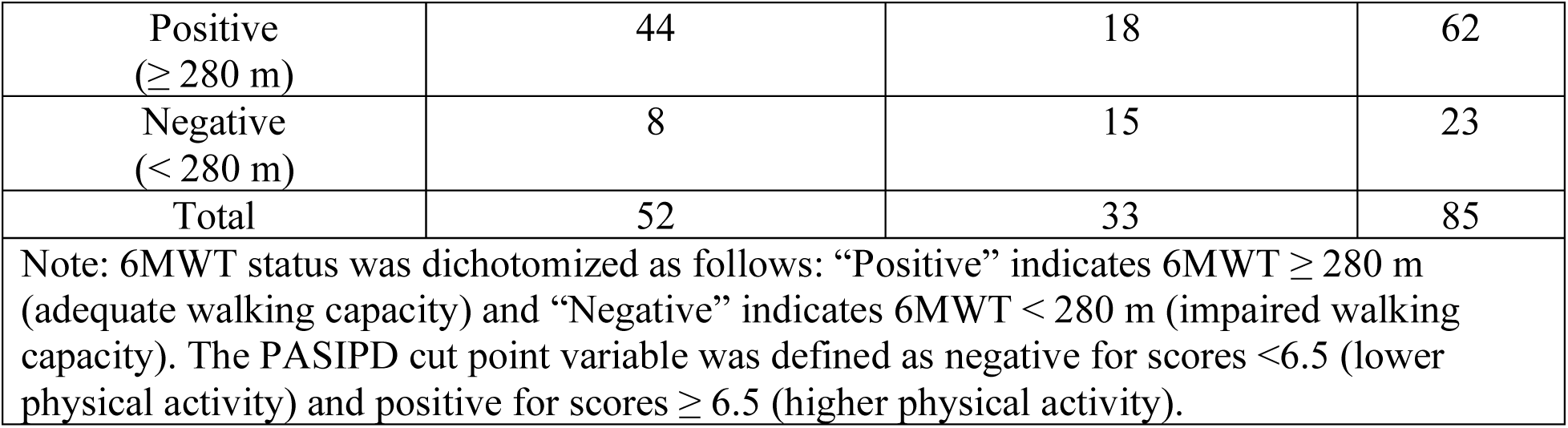
Contigency table for 6MWT Classification (≥280 m) and Youden-derived PASIPD cut point that maximizes the sum of sensitivy 1-specificity (≥ 6.5 MET hours/week)

**Table S12.** Contigency table for 6MWT Classification (≥380 m) and Youden-derived PASIPD cut point that maximizes the sum of sensitivy 1-specificity (≥ 10.6 MET hours/week)

| Table S12. Contingency table for 6MWT Classification ($\geq 380$ m) and Youden-derived PASIPD cut point that maximizes the sum of sensitivity 1-specificity ($\geq 10.6$ MET hours/week) | | | |
| --- | --- | --- | --- |
|  | PASIPD cut point |  |  |
| 6MWT cut point | Positive<br>($\geq 10.6$ MET hrs/wk) | Negative<br>($< 10.6$ MET hrs/wk) | Total |
| Positive<br>($\geq 380$ m) | 28 | 14 | 42 |
| Negative<br>($< 380$ m) | 8 | 35 | 43 |
| Total | 36 | 49 | 85 |
| Note: 6MWT status was dichotomized as follows: “Positive” indicates 6MWT $\geq 380$ m (adequate walking capacity) and “Negative” indicates 6MWT $< 380$ m (impaired walking capacity). The PASIPD cut point variable was defined as negative for scores $< 10.6$ (lower physical activity) and positive for scores $\geq 10.6$ (higher physical activity). | | | |

## Appendix 3

**Figure S1.**
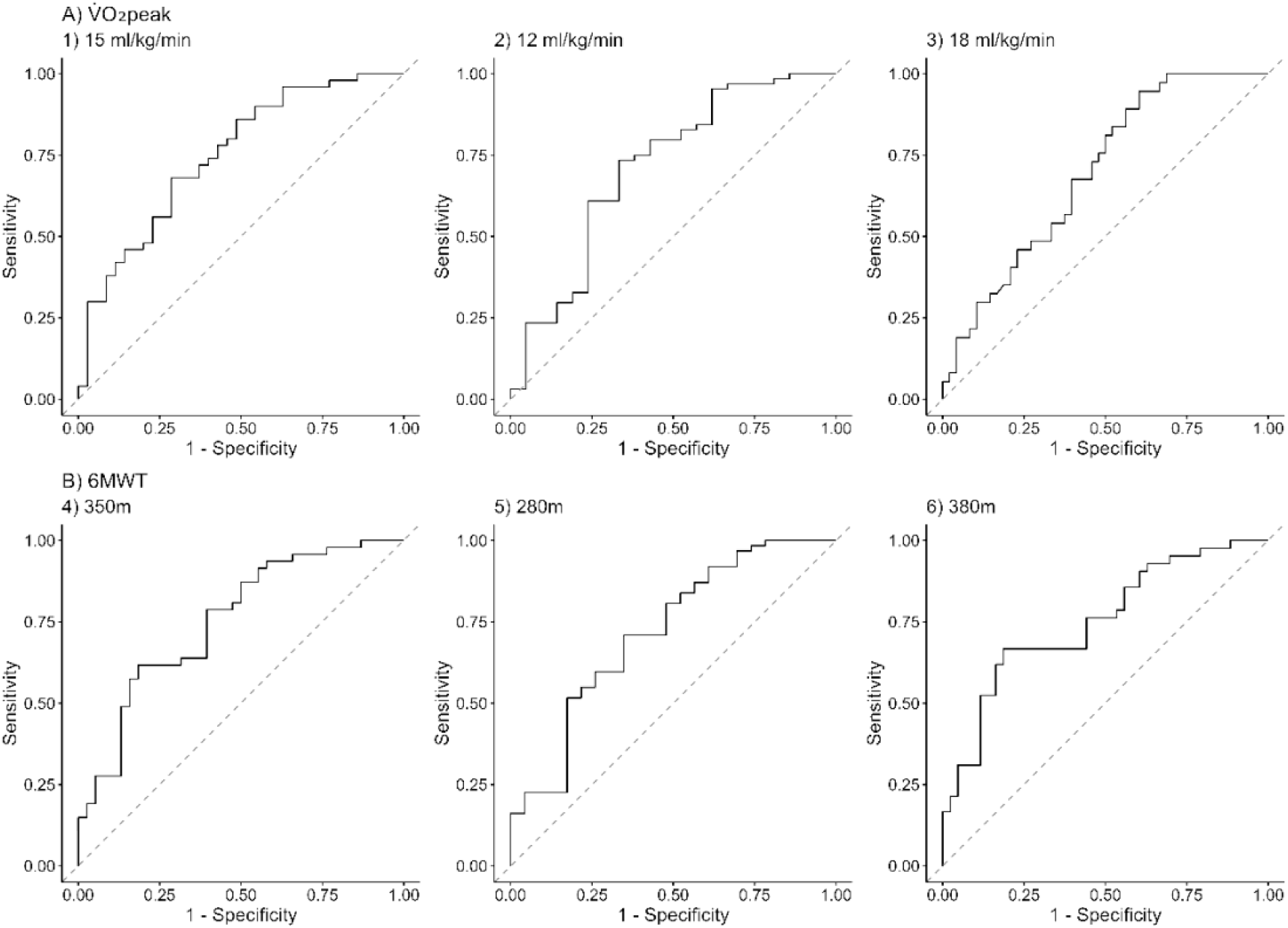
The unadjusted receiver operator characteristic curves for A) V̇O_2_peak cut points (Panel 1 to 3) and B) 6MWT cut points (Panel 4 to 6).

## Notes

### Competing Interest Statement

The authors have declared no competing interest.

### Clinical Protocols

https://clinicaltrials.gov/study/NCT03614585

### Author Declarations

Research ethics approval was obtained (Hamilton Integrated Research Ethics Board 4713, Centre de recherche interdisciplinaire en réadaptation du Montreal métropolitain-1310-0218) and informed written consent was obtained from all participants.

